# Transforming diagnostic capacity by syndromic infection testing for remote isolated communities

**DOI:** 10.1101/2024.11.11.24316818

**Authors:** Matthew Dryden, John Lee, Janice Toplass, Nicholas Cortes, Fabio Russo Del Piano, Tiffanie Skerritt, Mykala A S. St.Hill, Chester Crowie, Geoffrey Benjamin, Arlene A S. Siebs, Jonathan Smellie, Bill Hardy, Everette S. Duncan, Natalie Wright

## Abstract

**Objective:** Many of the UK overseas territories are small with limited microbiological diagnostic capacity for pathogens and antimicrobial resistance detection. The Covid-19 pandemic highlighted the particular vulnerability of these territories for the emergence of novel infections and antimicrobial resistance.

**Methods:** The UKHSA program with its territory partners implemented rapid automated syndromic molecular diagnostics (RASM), aiming to improve diagnostic capacity. Local laboratory staff were trained in the use of the diagnostics, and guidance was provided to clinicians for requesting tests. Syndromic diagnoses included enteric, respiratory, bloodstream, neurological infection, and antibiotic resistance mechanisms. Data on diagnostic capacity, turnaround times and clinical impact were collected from records before and after implementation of RASM.

**Results:** Turnaround time for results went from an average 14 days to 1 day, and often much shorter. Previously undiagnosed conditions, could now be identified to the microbiological level rapidly in territory, allowing appropriate specific clinical management, infection prevention, improved antimicrobial stewardship and rapid public health response. This technology is simple to operate and maintain with little scope for user error. The speed of microbiological diagnosis for patient management and public health detection and response was greatly enhanced.

**Conclusion:** Rapid microbiological diagnosis on site transformed patient management, the timely investigation and management of outbreaks and clusters, accurate surveillance and antimicrobial stewardship. Targeted RASM is cost effective, reducing the requirement for highly trained scientific staff and expensive logistics around rapid transport to reference laboratories. This innovation improves clinical care and strengthens local preparedness in communicable disease and public health response.

## Introduction

The COVID-19 pandemic highlighted the global inadequacy of infectious disease diagnostics and surveillance. Small island populations, such as the UK Overseas Territories (UKOTs), faced significant vulnerability due to limited laboratory capabilities and the delay in obtaining diagnostic results from distant reference facilities. Small, remote and isolated communities have always been highly vulnerable to the importation of infection and the lack of rapid on site microbiology diagnostics make appropriate rapid management of individual cases and clusters of infection a significant challenge. Low resource health systems are especially vulnerable to imported infectious disease and antibiotic resistance, largely due to inadequate laboratory diagnostics. ^1^

The UKOT Laboratory Strengthening Project focused on enhancing laboratory capacity during the pandemic, primarily through the implementation of open and closed platform RT-PCR for COVID-19 testing. However, the sustainability of open platform PCR testing in small laboratory settings was often limited, largely due to workforce skills and capacity.

These circumstances highlighted the vulnerability of the small, isolated communities to infectious disease in terms of diagnostic capability and surveillance. To address this the UKOT program developed a strategy of syndromic diagnosis development to enhance the range of diagnostic capacity locally and reduce the dependence on specialized reference laboratories with the logistical problems of sample transport and the delay in receiving a result which could be acted on. The UKOT programme facilitated the development of rapid automated syndromic molecular diagnostics (RASM) to detect multiple infectious disease targets from a single sample as a post-COVID legacy.

This study describes the process and clinical implications of implementing RASM in the UKOTs, evaluating the outcome in terms of enhancement of individual patient management, communicable disease control, and antimicrobial stewardship. The goal was to establish how RASM could support healthcare in small, resource-challenged territories by comparing before and after RASM implementation, looking at the following specific criteria:

1. The range of pathogens detected in clinical samples ie diagnostic capacity.
2. The clinical impact of diagnostics to individual patients based on case studies.
3. The detection of the causes of clusters and outbreaks.
4. Whether diagnostics could support public health management and infection prevention control.
5. Whether antibiotic resistance detection could support antimicrobial stewardship.

## Methods

The UKOT program is part of UK Health Security Agency (UKHSA) and has a specific remit and budget from the UK Foreign and Commonwealth Development Office (FCDO) to support health service development and in particular laboratory capacity for infectious disease and early recognition of outbreaks and emerging diseases. The UKOTs included in this study are broadly divided into the Caribbean region (Montserrat, Anguilla, Turks and Caicos Islands, Cayman Islands) and the Atlantic region (Gibraltar, St Helena, Falkland Islands, and Ascension).

In March 2023, the programme funded and delivered RASM through Biofire FilmArray TORCH equipment (Biomerieux), reagents, and training to eight UKOT laboratories, including Montserrat, Anguilla, British Virgin Islands, Turks and Caicos Islands, Cayman Islands, St Helena, Falkland Islands, and Ascension. The UKOT programme provided support in implementation, validation and laboratory governance. Other UKOTs such as Bermuda and Gibraltar had already developed molecular diagnostic capacity and purchased equipment themselves. Very small UKOTs such as Pitcairn Island and Tristan da Cunha were not included as they did not have diagnostic laboratories. Implementation took place between May and June 2023 and was achieved by on-site visits and on-line training. RASM panels included were enteric, respiratory, neurological and blood stream infections. The targets for these are listed in Table 1. The blood stream infection panel also included targets for the detection of antimicrobial resistance genes. Equipment was accommodated in the main hospital laboratory for each territory. In two UKOTs the equipment was accommodated in a nearby separate public health laboratory.

**Table 1.**
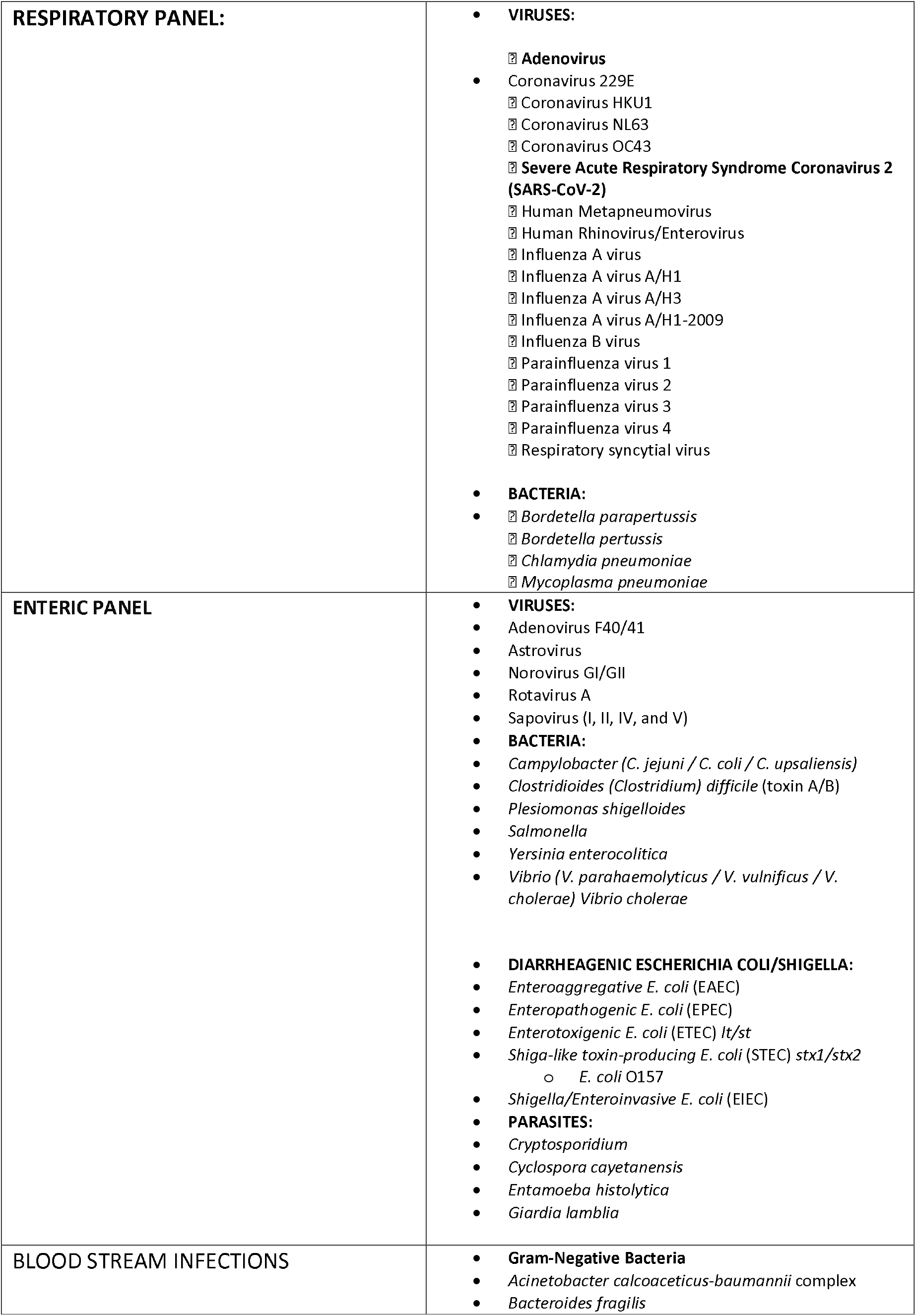
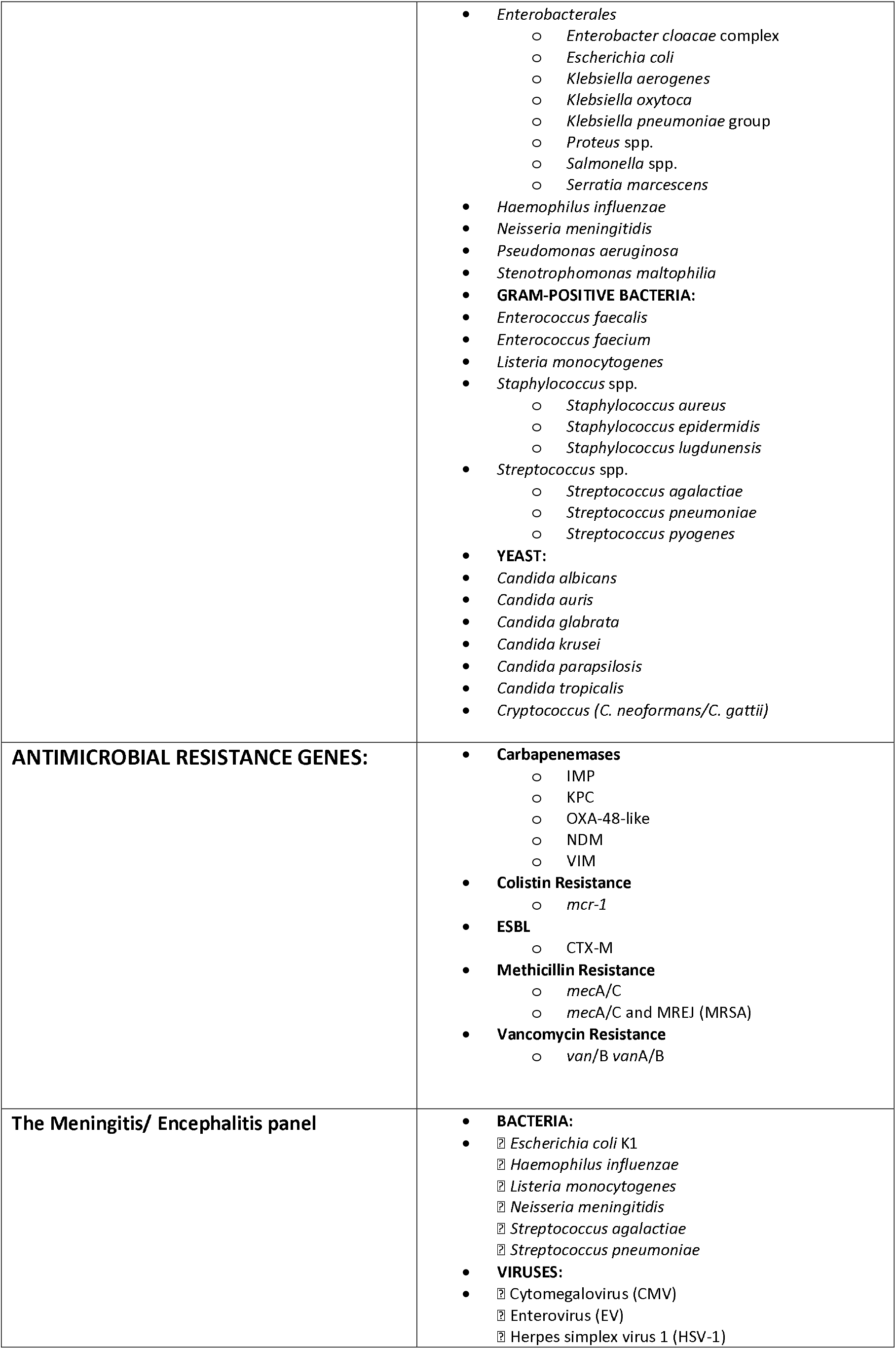
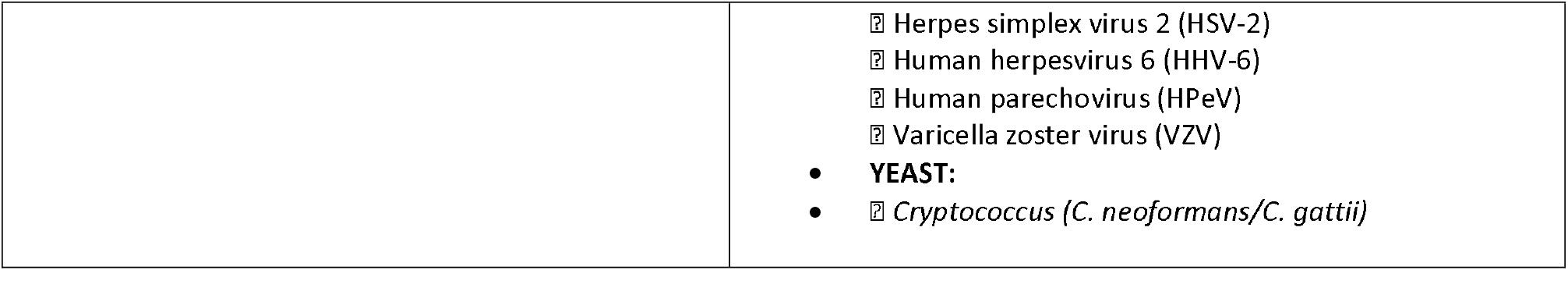
RASM targets.

Requestors of laboratory tests, mostly frontline clinicians in community facilities and hospital and public health personal were provided with a guide on the use and appropriate targeted requesting on samples to be tested by RASM.

Laboratory and/or infection prevention leads were asked to complete a brief survey for the year prior to syndromic diagnostic implementation. They were asked to collect data from laboratory workbooks, laboratory information systems (if they existed) and data on surveillance from the chief medical officer.

Specifically they were asked to collect the numbers of specific pathogens detected in the laboratory: enteric (Salmonella spp., Shigella spp., Campylobacter spp.), respiratory pathogens, blood stream infection, and antimicrobial resistant pathogens, specifically methicillin-resistant Staphylococcus aureus (MRSA), glycopeptide-resistant enterococci (GRE), extended spectrum beta-lactamase producing enterobacteriales (ESBL), and carbapenemase-producing bacteria (CRE). Turnaround times for laboratory results were also collected. Surveillance, outbreak and notifiable disease data was sought from the CMOs office.

Similar data were collected the year after implementation of syndromic diagnosis by RASM in June 2024. The range of pathogens detected was recorded and clinical examples of case and outbreak management were documented, including the time taken for crucial results to be available. In some similar cases it was possible to make a direct comparison of turnaround times and how this affected management.

The UKHSA Research Ethics Governance Group (REGG) granted ethical approval for this evaluation (NR0374), alongside a separate study examining the behavioural aspects of novel diagnostic use.

## Results

### Before RASM Implementation

In the year before implementation of RASM, two of the UKOTs had little or no phenotypic microbiology and so were unable to provide bacteriology or susceptibility testing. They were therefore unable to identify any cause of respiratory, enteric, bloodstream or neurological infection. The other UKOTs had microbiology services and were therefore able to identify most bacterial pathogens within the limits of routine culture technology and carry out susceptibility testing. Virology was extremely limited.

Several clusters or outbreaks of respiratory and enteric illness were suspected, but, as these could not be identified to the pathogen level, public health intervention was limited.

Complete pre and post RASM implementation data was available from 8/10 UKOTs. Of the two whose data was unavailable, one was slow in getting RASM established as a routine diagnostic service, the other RASM was only available in the private health sector and data was not available.

Diagnostic capacity in the pre-implementation phase is listed in Table 2. In the year prior to RASM implementation, 3/8 UKOTs identified respiratory infection outbreaks which were not caused by SARS Covid-19 (all UKOTs had implemented covid testing capacity during the pandemic). The cause of these outbreaks was not determined but presumed to be influenza. Two of the UKOTs identified enteric infection outbreaks. The microbiological cause and source was not determined.

**Table 2.** Summary of laboratory capacity prior to RASM implementation in the 8 UKOTs.

| **Diagnostic Capability** | **Caribbean 1** | **Caribbean 2** | **Caribbean 3** | **Caribbean 4** | **Atlantic 1** | **Atlantic 2** | **Atlantic 3** | **Atlantic 4** |
| --- | --- | --- | --- | --- | --- | --- | --- | --- |
| Respiratory pathogen detection | No | Limited bacteriology and virology | Limited bacteriology, no virology | Limited bacteriology, no virology | Limited bacteriology, no virology | Limited bacteriology, no virology | Limited bacteriology and virology | No |
| Enteric pathogen detection | No | Limited | Limited | Limited | Limited | Limited | Limited | No |
| Antimicrobial Susceptibility test | Limited | Phenotypic | Phenotypic | Phenotypic | Phenotypic | Phenotypic | Phenotypic | Not available |
| Turnaround time for blood cultures | 2-5 days | 2-5 days | 2-5 days | 2-5 days | 2-5 days | 2-5 days | 2-5 days | Not available |
| Turnaround time for reference tests | 2-4 weeks | 2-4 weeks | 2-4 weeks | 2-4 weeks | 4-8 weeks | 2-4 weeks | 2-4 weeks | 4-8 weeks |
| Outbreaks of enteric infection reported | 2 | 1 | 2 | 0 | 0 | 0 | 2 | 0 |
| Outbreak of respiratory infection reported | 1 | 0 | 0 | 0 | 1 | 0 | 1 | 1 |

Four UKOTs were unable to detect key antimicrobial resistance (AMR) determinants – methicillin resistance, glycopeptide resistance, extended spectrum beta lactamase (ESBL) production and carbapenem resistance (CRE).

### Post RASM implementation

Comparative data on the use and utility of RASM was available from 8 UKOTs one year after implementation. All UKOTs were able to detect a hugely expanded range of pathogens rapidly and on site in all the syndromes covered by the RASM panels – respiratory, enteric, bloodstream infection and neurological infection (Table 3). Rapid microbiological diagnosis allowed early appropriate patient management. It supported antimicrobial stewardship, ensuring appropriate starting or stopping of antibiotics and adjustment of antibiotic class. Rapid diagnosis permitted early public health intervention of clusters or outbreaks of disease, early infection prevention action and in the case of the single high-consequence infectious disease, the early diagnosis, described in vignette 4 below, allowed early activation of the cholera outbreak plan supporting rapid control of transmission and implementation of appropriate infection prevention in hospital and the community. It allowed early investigation of risk factors, sources of infection and routes of transmission. RASM enabled all of these actions in a timely manner, in a way which was not available to the UKOTs prior to RASM implementation.

**Table 3.**
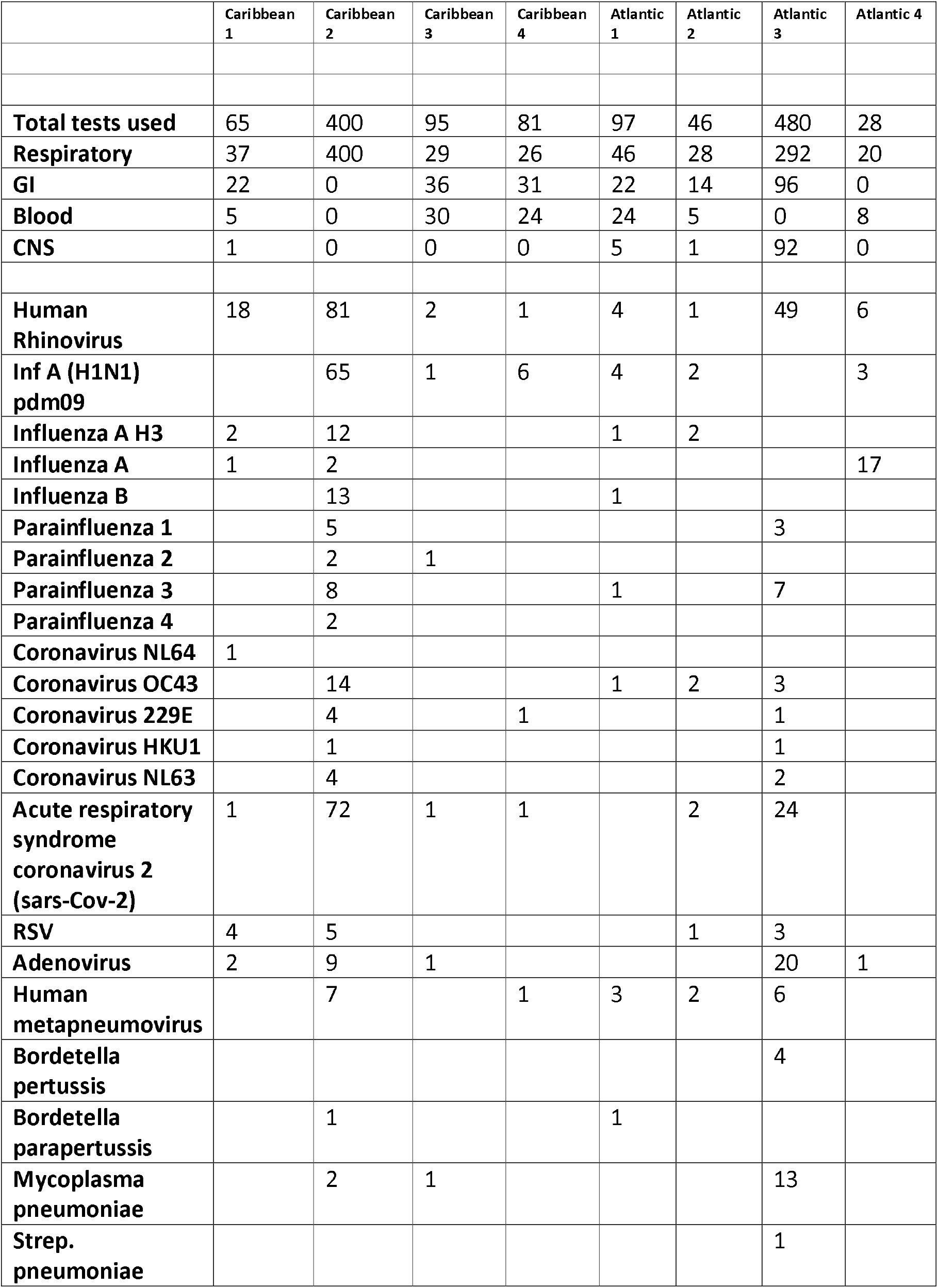
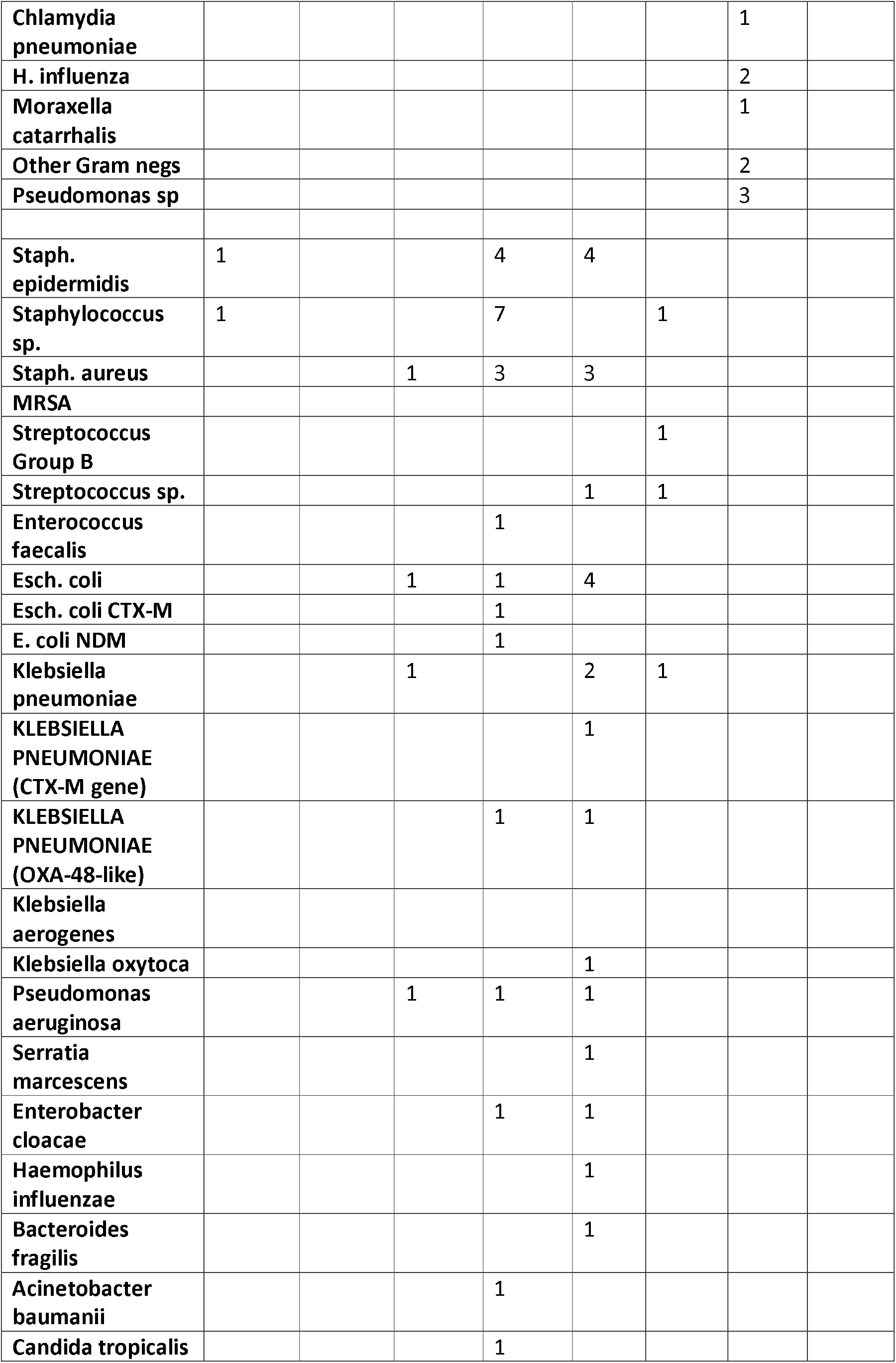
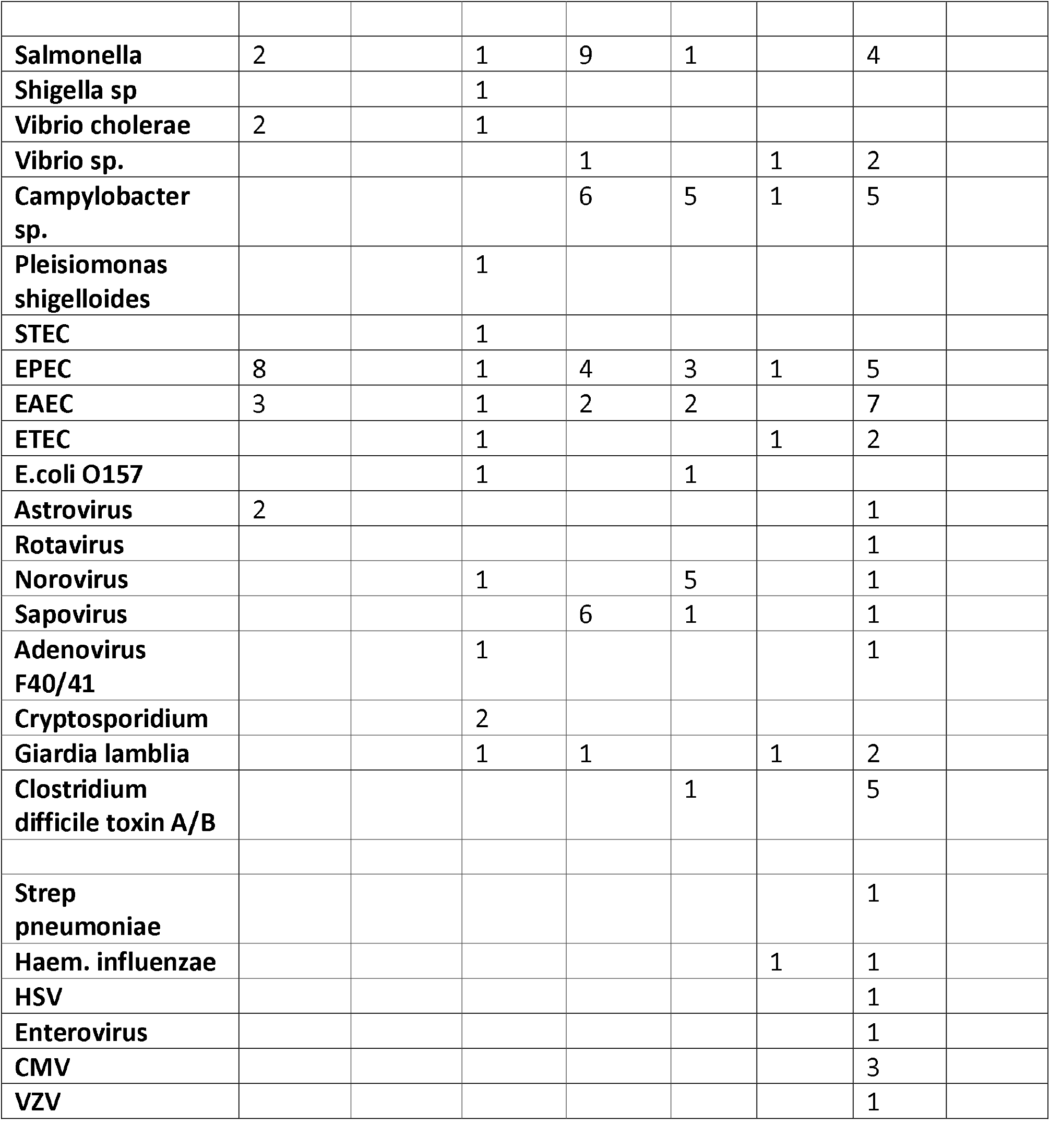
RASM usage and pathogen detection in the 8 UKOTs.

Guidance had been provided to the UKOTs on appropriate clinical requesting for the investigations. The siting of the equipment varied between UKOTs, some in hospital laboratories and some in public health laboratories. Because these laboratories had different functions, the diagnostic indications and use varied slightly from territory to territory. Some of the smaller UKOTs with populations of under 5000, had no neurological infection or in one case no means of performing blood cultures. The use of the RASM investigations and the pathogens detected are in Table 3.

Turnaround times for receiving clinically relevant results were dramatically shortened. Blood stream infections typically took 3-4 days to receive an isolate identification and antimicrobial resistant mechanism detection pre-RASM, compared with post-RASM 1 day in total or 90 minutes from a blood culture signalling positive to get an isolate identification and antibiotic resistance mechanism result. Subsequent antibiotic susceptibility testing was carried out phenotypically. Detection of respiratory and enteric pathogens went from unavailable pre-RASM to 90 minutes post RASM. For reference lab samples, results often took several weeks, but with the extended targets post RASM, an answer was often available in 90 minutes.

The effect of providing rapid diagnostics delivered by laboratory technologists is best illustrated by clinical vignettes, five of which are listed below.

### Public health intervention in outbreaks

1. **Community respiratory infection cluster:** patients presented to a clinic with acute respiratory illness. Patients were clinically stable and hospital admission not required except for one infant for observation overnight. There was public health concern that this may be a new wave of Covid-19. Nasopharyngeal swabs were sent urgently to the laboratory. Samples were processed on RASM respiratory panel. The result was communicated to the clinical team within 90 minutes demonstrating that this cluster was caused by Metapneumovirus. The result also supported antimicrobial stewardship as no antibiotics were required for patients.
2. **Community enteric outbreak** – Early assessment suggested more than 10 cases were affected with acute diarrhoea. All had attended an extended community event where locally produced food and fast food was served. One elderly patient with diabetes and heart failure was admitted to hospital with dehydration. Stool samples were sent to the laboratory from the hospitalised patient. RASM enteric panel identified Salmonella sp. food poisoning type in 90 minutes. Other samples confirmed the cause of the outbreak. Public health control was implemented rapidly. Antibiotics was given to the hospitalised patient, but no other antibiotics required.

### Management of individual cases and antibiotic stewardship

3. **Blood stream infection and susceptibility testing:** A patient was admitted to hospital with sepsis. She had had recent biliary surgery abroad. The admitting diagnosis was suspected biliary sepsis. The patient was resuscitated and received ionotropic support. Empirical antibiotics – piperacillin-tazobactam were prescribed. Blood cultures collected on admission signalled positive in 8 hours showing Gram negative rods. Blood was subcultured but also run on RASM bloodstream infection panel. The result was available in 90 minutes; Klebsiella sp was identified and extended spectrum beta lactamase production was detected. Antibiotics were appropriately switched to meropenem.
4. **Meningitis:** An adult presented with fever and seizures and signs of meningitis. Lumbar puncture demonstrated meningeal inflammation with a raised cerebrospinal white count. CSF was analysed on RASM neurological panel. At 40 minutes, Strep. pneumoniae was detected. Antibiotics were rationalised. Antiviral was stopped. Widespread antibiotic prophylaxis for contacts was avoided.

### Managing High Consequence infection

5. A traveller in the region returned to home territory with watery diarrhoea, vomiting and collapse from dehydration. The patient was resuscitated in the emergency department and admitted to hospital in isolation. Stool sample was sent for RASM enteric panel with a result available in 90 minutes. Vibrio cholerae O1 was detected. The territory cholera plan was rapidly implemented, a public health incident group was called and public health measures to prevent further spread and investigate other possible cases implemented rapidly.
6. **The effect on health service resources in relation to suspected Middle Eastern Respiratory Syndrome (MERS) before and after RASM**

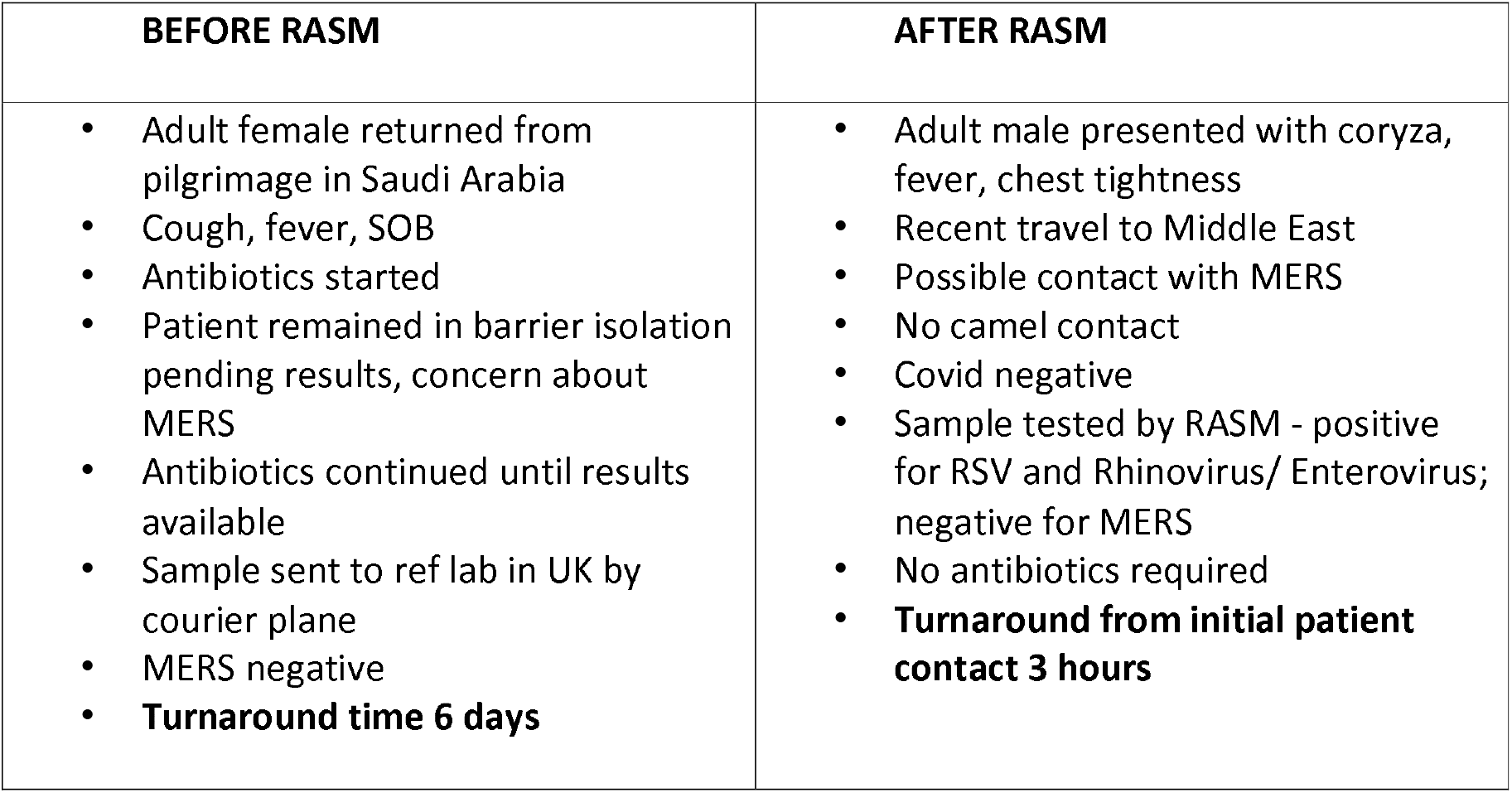

Both patients had suspected Middle Eastern Respiratory Syndrome (MERS). Before RASM the patient required prolonged isolation, antibiotics, and expensive transportation of a sample to a reference laboratory with a long turnaround time. After RASM, MERS was excluded with the on site test within 3 hours of initial contact with the patient.

## Discussion

RASM has proved to be transformative for several UKOT laboratories that lacked the technology, workforce, and diagnostic capacity to identify a broad range of pathogens. Low-resource settings are disproportionately burdened by infectious diseases and antimicrobial resistance.^1^ Accurate microbiological diagnosis is essential for appropriate individual patient management and effect public health control of infection incidents, clusters and outbreaks. Without clear diagnosis, clinical management is at best a guess, there is a waste of resources, there is inappropriate use of antibiotics and public health control relies on intuition alone. Failure to deliver a clear diagnosis contributes to morbidity and mortality. The recent SARS-Covid-19 pandemic has highlighted the need for early warning systems to detect emerging infection and its spread, and RASM has clearly extended this capacity in a way which was impossible before. While the pandemic and encroaching AMR has highlighted, like never before, the necessity for improving diagnostic capacity, it is not enough to simply supply equipment and reagents. Laboratory governance has to be at the heart of such developments.^2^

The UKOTs included in this study vary significantly, not least in population size from 800 (Ascension) to 68,000 (Cayman). They all have self-determining governance which includes responsibility for health services. Most of the UKOTs are small, remote and isolated and have limited workforce capacity. All these characteristics make it challenging to develop microbiology services which deliver timely and responsive results. The UKOT programme funded by FCDO was set up during the Covid pandemic to support the UKOTs in public health and developing diagnostic capacity. During the pandemic, open platform PCR was set up in each of the UKOTs to detect Covid-19. Open platform PCR investigations require specialist technical skills and these are not sustainable in some of the UKOTs where laboratory scientists are mostly generalists with a wide range of responsibilities in their laboratories. It is simply not possible to have the workforce capacity to have broad open platform molecular laboratory in each UKOT.

Therefore in 2022, at the end of the pandemic, the UKOT programme looked to alternative solutions for enhancing diagnostic range and capacity and develop an early warning system for infection within the territories. Previously any samples for complex investigation had to be sent to different reference laboratories around the world. The difficulty of transport logistics, cost and turn around times limited the efficacy of these reference services. Furthermore, during periods of lockdown & travel restrictions such as those implemented during the COVID-19 pandemic further reduction in flights and shipping traffic increased UKOT isolation and compounded logistic challenges, further emphasising the need for within territory diagnostic resilience.

A strategy of syndromic diagnosis was chosen and the most appropriate commercial equipment to deliver this was the Biofire filmarray technology. In addition, Cepheid GeneXpert technology was also considered for other important areas of diagnostic need, notably sexually transmitted diseases, HIV viral load, hepatitis serology, human papilloma virus, tuberculosis. These technologies allowed non-specialist scientists to operate the equipment and deliver rapid investigation results in territory. The UKOT programme sought the funding to purchase and implement these technologies and successfully achieved this my mid 2023. This study has reported the impact of delivering rapid diagnostics to these remote locations.

This automated syndromic testing approach proved to be transformative for several UKOT laboratories that lacked the technology, workforce, and diagnostic capacity to identify a broad range of pathogens. It has also provided the UKOTs the ability to diagnose infection quickly, a key advantage in the management of individual patients and controlling infection.

The advantages of RASM have been clear, ensuring improved clinical outcomes, biosecurity, and response capabilities. Individual patient management is compromised unless there is a clear microbiological diagnosis. With a rapid diagnosis, the most appropriate antibiotics can be administered quickly, or antibiotics stopped quickly if they are not indicated. Early diagnosis has supported antimicrobial stewardship and limited resistance selection. Early diagnosis also supported appropriate infection prevention intervention within healthcare settings.

Syndromic diagnosis has allowed early public health intervention of respiratory and enteric clusters and outbreaks for the first time and has improved the accuracy and quality of surveillance. The early diagnosis of cholera and exclusion of MERS coronavirus in individual cases resulted in a rapid public health response in the first and a scaling down of resource intensive interventions in the second.

This study has not carried out a cost effectiveness analysis. Molecular diagnostics, including RASM, are costly and the consumables have to be purchased and be available with an adequate shelf life in a timely way. Supply lines can be complex in remote territories. Nevertheless, targeted RASM is likely to be cost effective for the UKOTs in comparison with the alternative costs of developing multiple diagnostic platforms with associated highly trained, specialist scientific staff, cost of transport of samples to reference facilities, cost of delayed public health or infection control intervention, prolonged isolation of patients, cost od collateral damage from inappropriate antimicrobials. Health ministries should be aware that the financing of rapid diagnostics is likely to be highly cost effective.^3^

It is difficult to manage outbreaks without good diagnostic data. Surveillance of infectious disease and antibiotic resistance has been limited across the UKOTs. Certain regions, especially the Caribbean, are susceptible to outbreaks because of increasing global travel. Prior to and since the Covid-19 pandemic the Caribbean has suffered epidemics of zika, dengue and chickungunya. Many other emerging and vector borne diseases could have major consequences for the UKOTs. Some UKOTs are at the crossroads of shipping lanes and have had incidents of potential introduction of high-consequence infections. Without rapid diagnostics, the risk to public health is significant. While current syndromic panels are unable to detect many causes of high consequence infection or emerging global diseases, the UKOT programme is working with all the territories to increase the range of diagnostics capability in this area of global infection by similar RASM technology. The programme strategy is to have local rapid diagnostics for most emerging and high-consequence infection within the next two years.

## Data Availability

All data produced in the present work are contained in the manuscript

## Acknowledgements and Declarations

The Authors would like to thank the Chief Medical Officers of the UK Overseas Territories for their wholehearted support of the study. The study had no specific funding but the authors acknowledge the generous funding from the UK Foreign and Commonwealth Office to develop diagnostic capacity in the UKOTs. The authors have no conflict of interest. The UKHSA Research Ethics Governance Group (REGG) granted ethical approval for this evaluation (NR0374), alongside a separate study examining the behavioural aspects of novel diagnostic use.

## Declarations

The authors have no declarations to make. The equipment and delivery of RASM was funded by the UK government, Foreign and Commonwealth Office via the UKOT programme of the UK Health Security Agency

